# Randomized Controlled Trial Effect of Short-Term MIND Diet Intervention on Hunger Hormones, Anthropometric Parameters, and Brain Structures in Middle-Aged Obese Women

**DOI:** 10.1101/2020.06.28.20142018

**Authors:** Golnaz Arjmand, Mojtaba Abbas-Zadeh, Majid fardaei, Mohammad Hassan Eftekhari

## Abstract

The growing prevalence of obesity and its destructive effects on the hormone’s circulation and brain structures has attracted attention to specific dietary patterns. This trial aimed to examine the effect of short-term newly Mediterranean-DASH Intervention for Neurodegenerative Delay (MIND) pattern on anthropometric parameters, hunger hormones as well as brain structures in obese women. The study was a randomized restricted controlled trial in which we analyzed 37 healthy obese women with a mean age of 48 ± 5.38 years and mean Body Mass Index (BMI) 32 ± 4.31 kg/m^2^. Participants were randomly allocated to a hypocaloric modified MIND diet or a hypocaloric waiting list control diet and followed the protocols for three months. Differences in anthropometric and laboratory analysis, as well as brain structures, were determined at baseline and follow up. There was a more significant reduction in weight, BMI, percentage of body fat, waist circumference, and Leptin concentration in the MIND diet group compared to the control group (*ps*<0.05). Despite a deceased in plasma levels of Ghrelin and GLP-1 in the control group, results have found a significant increase in these hunger hormones in the MIND diet group (*ps*<0.05). Our results diminished to detect any differences in the whole and regional brain structures between two groups after three months follow up. Our trial for the first time demonstrated that a three-month MIND diet intervention could improve the potentially devastating effect of obesity on metabolic profiles and anthropometric parameters. However, we couldn’t find the MIND diet effect on brain structures.

## 1 Introduction

Obesity and overweight are accompanied by an alteration in body composition, impair hormonal secretion and inefficiency in a network between the endocrine and nervous systems [1]. Despite extensive effort to control obesity, according to 2020 WHO reports, worldwide obesity has nearly tripled since 1975.

Concerning the endocrine system, some hunger hormones, such as Leptin, Ghrelin, and GLP-1, has an essential role in conveying information about the energy stores to hypothalamus centers. Some evidence exists to support the idea that Leptin resistance, which is happening in obesity, blunt its effect in the hypothalamus and increases body weight gain. Additionally, obesity desensitizes Ghrelin cells to signals of caloric restriction and food intake, which can attenuate weight gain and the development of glucose intolerance [2]. Besides, recent studies have shown that diurnal secretion of GLP-1 was dramatically decreased in morbid obesity [3]. This indicates that unique dietary patterns and weight loss can improve its secretion.

Brian’s structural changes in obesity are less well known. However, previous studies provide evidence that higher body mass index alters gray matter and white matter volume of some brain regions [4]. A voxel-based morphometry structural study found that BMI in obese subjects negatively associated with gray matter volume of middle frontal and postcentral gyrus [5]. Additionally, obesity amplifies the risk for dementia even in healthy individuals by deteriorating cerebral atrophy and increase their vulnerability to future neurodegenerative diseases [6].

In this regard, evidence has shown that dietary-induced weight loss improves the metabolic profile and brain structures in obese patients [7]. Published studies agree in this statement that improvement occurs before a significant amount of weight is lost [8]. Previous studies have examined the effect of healthy eating patterns, including the DASH diet as well as the Mediterranean diet, with obesity, but these findings are inconsistent [9]. A new dietary MIND pattern was designed by Morris and her colleagues in 2015. The MIND diet has been recognized from comprehensive epidemiologic studies as one of the healthiest diets. However, the MIND pattern is based on the Mediterranean and the DASH diet and shares many food groups with them for neuronal protection use. It differs by allocating separate categories for the consumption of green leafy vegetables, berries, and olive oil as brain-healthy foods and red meat, sweets and fast foods as unhealthy brain foods that have been reported in neuroprotective features [10].

Given the growing prevalence of obesity, and detrimental impact on general and brain health, the need for interventions that can reverse this vicious cycle is increased. Since the MIND dietary pattern was based on the Mediterranean and DASH diet, it can be concluded that, in addition to having benefits for brain health, the MIND diet might also be useful in weight loss and general health. In addition, obesity in mid-life can be considered as a chronic and significant risk factor for global death in the future. From what we know, there was not any Randomized Controlled Trial (RCT) to assess the effect of the MIND diet on the component of obesity. Therefore, we designed a randomized trial in middle-aged women to investigate the effect of the short-term intervention of MIND dietary pattern on anthropometric parameters and hunger hormones in healthy obese women along with brain structure investigation.

## 2 Methods

### 2.1. Study overview

The study was a three month, randomized, single-blind, two-parallel arms trial; we determined the effect of the MIND diet on changes in anthropometric and metabolic profiles as well as brain structure in middle-aged obese women.

#### Participants

Forty healthy obese middle-aged (40-60 rears) women with BMI 30-35 kg/m^2^ have entered the study through advertising recall from Imam Reza Clinic of the Shiraz University of Medical Science located in the center of Shiraz, Iran. The eligibility criteria were: Mini-Mental State Examination survey (MMSE) ≥ 24, no history of severe medical problems, participation in weight loss programs, or using weight loss drugs in the last three months. Initial eligibility of participants was first asked during a telephone screening. We also excluded people if study protocols were not entirely followed by them or become pregnant.

Participant’s recruitment was performed after approval by the Ethics Committee of the Shiraz University of Medical Science, and participants provided signed informed consent before participation. The study was carried out in agreement with the principles of the Helsinki Declaration.

### 2.2. Study protocol

After admitting potentially eligible participants to the study, individuals were randomly allocated to the calorie-restricted MIND diet and calorie-restricted waiting list control diet. The allocation sequence was generated by a computerized random number generator. At the first visit, the dietary history of individuals was assessed using a 168-item semi-quantitative food frequency questionnaire (FFQ), and energy needs were calculated using World Health Organization equations [11] to estimate basal metabolic rate. Meals in both calorie-restricted diet groups included a balanced mix of foods with 50-55% carbohydrates, 30% fat, and 15-20% proteins, and we restricted calorie intake to at least 1500 kcal/day. In the MIND diet group, participants are trained to modify their dietary patterns towards the MIND diet instruction for a three months study period. The mentioned diet was developed to recommend further the consumption of green leafy vegetables, berries, nuts, fruits, and legumes (especially beans). The diet also emphasizes olive oil as the primary source of fat in the diet. In the MIND diet, it is also recommended to eat fish and poultry more than one meal and more than two meals a week, respectively. Also, in the current diet, to improve brain function and general health, the consumption of red meat, butter, and all kinds of margarine, as well as prepared foods and sweets, has been limited. All of the recommended components were based on energy needs for each person. The rationale for using the calorie restriction for both groups was to enable assessing the effect of the only MIND diet as the primary intervention and for ethical issues. The control group was told not to change their usual eating habits during the three months, and they were instructed to have a calorie-restricted diet alone. Additionally, at the end of the study, the MIND dietary recommendation was given to the waiting list control group. The participants received a menu based on groups recommendation weekly. Adherence to the MIND diet was monitored by MIND diet score questioner (Table 2), shopping list [10] as well as a three-day food diary.

### 2.3. Blood sampling, Clinical and Anthropometric Data

After overnight fasting, the blood sample was collected from participants at the beginning and end of the three months. The blood samples were centrifuged at 5,000 rpm for 5 min, and their isolated serums were placed inside micro-tubes. The collected samples were then stored at −80°^c^ for further analysis. To determine plasma levels of Leptin, Ghrelin, and Glucagon-like peptide-1(GLP-1), we used Enzyme-linked Immunosorbent Assay (ELISA) specialized kits (Bioassay Technology Laboratory) before and after three months of study according to the manufactured instruction. All of the laboratory measurements were done in the same laboratory.

Furthermore, weight, height, waist circumference, as well as the percent of body fat were measured at all 2-time points of study. Body weight was measured using a digital scale to the nearest 0.1 kg, and standing height was measured using a stadiometer to the nearest 0.1 cm. Body composition was obtained with a Bioelectrical Impedance Analysis device (BIA) to the closest according to a standardized protocol (In Body S-10, USA). We determined total body water, percent of body fat, fat-free mass, and weight by using BIA. Hip circumference was measured by using a plastic fiber tape placed directly on the skin at the Iliac crest and maximum extension of the buttocks. Participants were directed not to engage in strenuous physical activity for 24 hours before the measurements and also not to drink alcohol or caffeinated beverages. Finally, waist circumference was measured at the narrowest part of the torso between the costal margin and Iliac crest.

### 2.4. MRI accusation

MRI scans were obtained by a 12-channel head coil 3T Siemens Skyra system on a subgroup of 11 participants in each group. T1-weighted images consisted of 192 slices and were used in the sagittal plane, which prepared with gradient-echo sequence (repetition time = 1900ms, echo time = 2.52ms, flip angle = 9°, voxel size = 1×1×mm). Image preprocessing was performed by the Freesurfer, stable version 5.1 (http://surfer.nmr.mgh.harvard.edu). This procedure included motion correction, intensity normalization, Talairach registration, skull stripping, segmentation of subcortical white matter, tessellation of the G.M./white matter (W.M.) boundary, automated topology correction, and surface deformation.

To reduce biased analysis and measurement noise and to record changes in brain structure in response to MIND dietary intervention in study groups, Freesurfer’s longitudinal processing method was used. For this method, images from the base and three months are first processed independently. Then, the baseline data were separated from the quarterly data in a vertex-vise manner. For group analysis, the output map for each participant was normalized to the common space (fsaverage) and flattened to the full width of half the maximum (FWHM) of 10 mm.

Freesurfer reconstructs white and gray matter surfaces. White matter surface or white surface is a boundary of white matter, and gray matter and gray matter surface or pial surface is the boundary of gray matter and pial. The spatial resolution of reconstructed surfaces is not restricted to voxel resolution of original images so that the surfaces can show the submillimeter differences. The distance between white and pial surfaces is defined as the cortical thickness, and it was calculated for each vertex. The cortical surface area was computed as the mean area related to the triangular region of each vertex. The product of cortical thickness and surface area for each vertex was calculated as the cortical volume. Furthermore, some anatomic parcellation of the cortex, which was performed by Freesurfer, was selected based on previous studies as a Region Of Interest (ROI) for region analysis [12].

### 2.5. Statistical Analysis

The primary endpoint of the study was described as the improvement in anthropometric and metabolic profiles of the MIND diet group after three months of research. The secondary endpoint was measuring differences in brain structure following the study. Data were analyzed using SPSS 22.0 (SPSS, Chicago, IL), and all data were presented as mean ± standard error. The normality of the data was confirmed using a one-sample Kolmogorov-Smirnov test. We also used the Mann-Whitney U test to compare the brain structure changes in the whole brain and ROI analyses between two groups. To show the effect of three months MIND diet intervention on variables, we run a mixed model two-way repeated-measures analysis of variance (ANOVA), where time (baseline, 3month) as a within-subject factor and treatment (MIND diet group, control group) as a between-subject factor.

*P*-value <0.05 was considered as a significant level for all analyzes, and corresponding effect sizes in the form of partial Eta square were calculated to evaluate the magnitude of effect for all hypotheses.

## 3 Results

### 3.1. Baseline characteristics

Between October 2018 and March 2019, 50 women were assessed for eligibility at one of the health clinics of the Shiraz University of Medical Science. As reported in the CONSORT flowchart diagram (Figure 1), thirty-seven healthy obese women with an average age of 48 ± 5.3 years were thoroughly followed the study protocols. There were no significant differences between the two group baseline characteristics (*p*>0.05), and no participant reported any adverse effect of interventions. Descriptive results showed that at baseline, the mean BMI of participants was 32.01± 4.21, and mean weight was 82.10±12.71. Most of the participants were married (83.80%), and the mean years of education in them was 16.4 ± 1.03 year. The two groups were comparable in terms of the baseline plasma Leptin levels. The baseline characteristic was shown in Table 1.

**Table 1.**
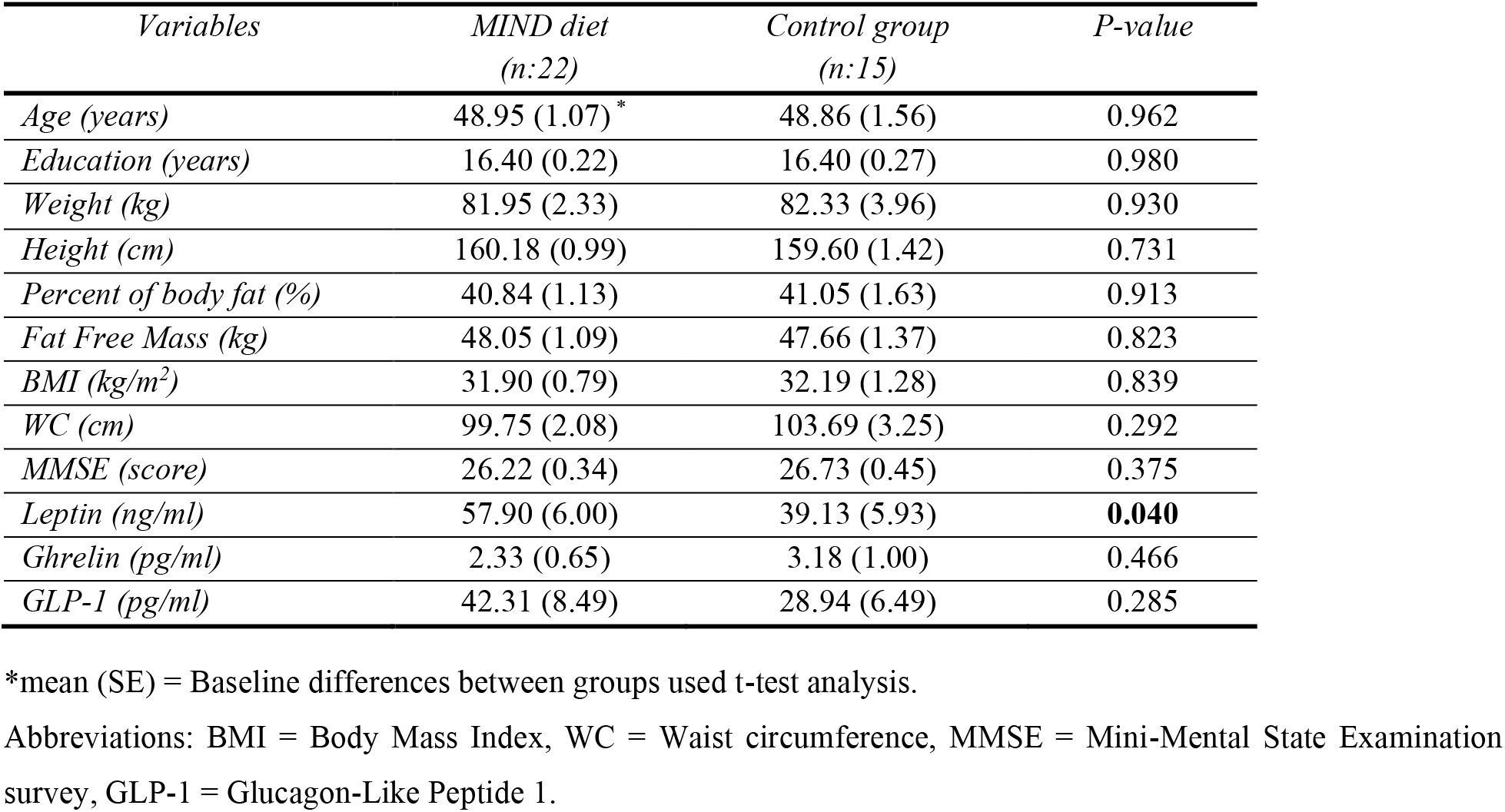
Baseline characteristics of participants in the MIND diet versus control group.

| *Variables* | *MIND diet*<br>*(n:22)* | *Control group*<br>*(n:15)* | *P-value* |
| --- | --- | --- | --- |
| *Age (years)* | 48.95 (1.07) * | 48.86 (1.56) | 0.962 |
| *Education (years)* | 16.40 (0.22) | 16.40 (0.27) | 0.980 |
| *Weight (kg)* | 81.95 (2.33) | 82.33 (3.96) | 0.930 |
| *Height (cm)* | 160.18 (0.99) | 159.60 (1.42) | 0.731 |
| *Percent of body fat (%)* | 40.84 (1.13) | 41.05 (1.63) | 0.913 |
| *Fat Free Mass (kg)* | 48.05 (1.09) | 47.66 (1.37) | 0.823 |
| *BMI (kg/m<sup>2</sup>)* | 31.90 (0.79) | 32.19 (1.28) | 0.839 |
| *WC (cm)* | 99.75 (2.08) | 103.69 (3.25) | 0.292 |
| *MMSE (score)* | 26.22 (0.34) | 26.73 (0.45) | 0.375 |
| *Leptin (ng/ml)* | 57.90 (6.00) | 39.13 (5.93) | **0.040** |
| *Ghrelin (pg/ml)* | 2.33 (0.65) | 3.18 (1.00) | 0.466 |
| *GLP-1 (pg/ml)* | 42.31 (8.49) | 28.94 (6.49) | 0.285 |

\*mean (SE) = Baseline differences between groups used t-test analysis.
Abbreviations: BMI = Body Mass Index, WC = Waist circumference, MMSE = Mini-Mental State Examination survey, GLP-1 = Glucagon-Like Peptide 1.

**Table 2.** Changes in food intake from baseline to follow up in the MIND diet group vs. control group.

| *MIND diet group (n:22)* |  |  |  | *Control group (n:15)* |  |  |
| --- | --- | --- | --- | --- | --- | --- |
| *Foods* | *Baseline* | *Follow up* | *P-value* | *Baseline* | *Follow up* | *P-value* |
| *Green leafy vegetables (serving/week)<sup>a</sup>* | 3.72±0.55 | 5.50±0.51 ▲ | < **0.001** | 3.60±0.50 | 3.93±0.25 ▲ | < **0.019** |
| *Other vegetables (serving/week)<sup>b</sup>* | 3.86±0.63 | 6.90±0.52 ▲ | < **0.001** | 3.93±0.25 | 4.00±0.65 ▲ | < 0.670 |
| *Berries (serving/week)* | 0.90±0.42 | 2.00±0.00 ▲ | < **0.001** | 0.93±0.25 | 0.73±0.59 ▼ | < 0.189 |
| *Nuts (serving/month)* | 4.45±1.01 | 4.63±0.49 ▲ | 0.478 | 4.53±0.63 | 3.73±0.59 ▼ | < **0.001** |
| *Olive oil (primary oil) \** | 0 | 1 | < **0.001** | 0 | 0 | 1 |
| *Butter, margarine (table spoon/day)* | 1.50±0.59 | 0.63±0.49 ▼ | < **0.001** | 1.20±0.41 | 0.46±0.51 ▼ | < **0.001** |
| *Cheese (servings/week)* | 6.40±0.50 | 1.68±0.56 ▼ | < **0.001** | 6.33±0.48 | 6.33±0.48 | < **0.001** |
| *Whole grains (serving/day)* | 1.00±0.00 | 2.18±0.39 ▲ | < **0.001** | 1.00±0.00 | 0.33±0.48 ▼ | < **0.001** |
| *Fish (not fried) (meals/month)* | 1.22±0.42 | 2.45±0.50 ▲ | < **0.001** | 1.33±0.48 | 1.00±0.00 ▼ | < **0.019** |
| *Beans (meal/week)<sup>c</sup>* | 1.36±0.49 | 2.92±0.52 ▲ | < **0.001** | 1.46±0.51 | 1.60±0.50 ▲ | < 0.164 |
| *Poultry (not fried) (meal/week)* | 2.09±0.29 | 2.86±0.35 ▲ | < **0.001** | 2.13±0.35 | 2.00±0.37 ▼ | < 0.164 |
| *Red meat and products (meals/week)* | 2.36±0.49 | 2.00±0.00 ▼ | **0.002** | 2.46±0.51 | 2.40±0.50 ▼ | 0.334 |
| *Fast fried foods (times/week)* | 1.18±0.39 | 0.45±0.50 ▼ | < **0.001** | 1.26±0.45 | 0.93±0.23 ▼ | **0.019** |
| *Pastries and sweets (servings/week)* | 4.77±0.68 | 2.50±0.51 ▼ | < **0.001** | 4.73±0.79 | 2.60±0.50 ▼ | < **0.001** |

<sup>a</sup>Kale, collards, greens; spinach; lettuce/tossed salad. <sup>b</sup>Green/red peppers, squash, cooked carrots, raw carrots, broccoli, celery, potatoes, peas or lima beans, tomatoes, tomato sauce, string beans, beets, corn, zucchini/summer squash/eggplant, coleslaw, potato salad. <sup>c</sup>Beans, lentils, soybeans.
Calculated using MIND diet score questioner and three-day food recall, completed four times using pair sample t-test and \* using Man-Whitney t-test.

**Figure 1.**
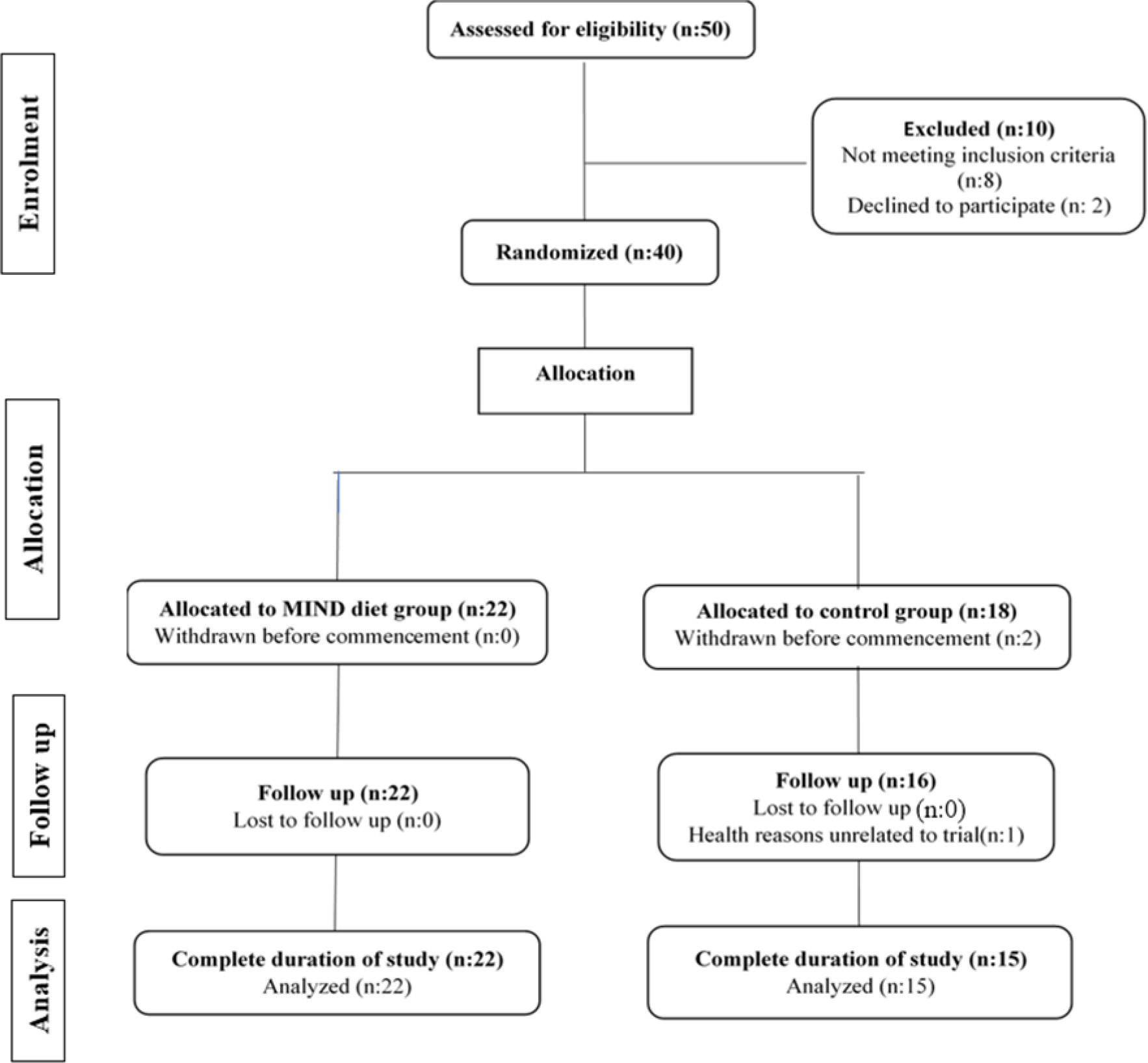
CONSORT flow diagram of the study.

### 3.2. Changes in food intake from baseline to follow up

As table 2. shown, the consumption of green leafy vegetables, other vegetables, berries, olive oil, fish, beans, and poultry significantly increased in participants in the MIND diet group (*ps*<0.05). The MIND diet group also considerably decreased the intake of butter, cheese, red meats, fast foods, and sweets (*ps*<0.05). There were no significant changes in nut consumption in the MIND diet group before and after three months.

### 3.3. Changes in Anthropometric Parameters

Mean changes in anthropometric data at baseline and three months follow up were summarized in Figure 2. The linear mixed model revealed a significant group × time interaction for body mass index (*p* = <0.001, Figure2, panel E) and weight (*p* = <0.001, Figure2, panel F), indicating better weight loss after three-month intervention in the MIND diet group in comparison with the control group. However there was no significant effect of time on total body water (*p* = <0.647, Figure2, panel A) or fat free mass (*p* = <0.486, Figure2, panel D); the three month MIND diet intervention had a statistically significant effects on percent of body fat (*p* = <0.032, Figure2, panel C) and waist circumference (*p* = <0.014, Figure2, panel G). Finally, it is indicated that the MIND diet has been able to reduce the percentage of body fat while maintaining whole-body water and also fat-free mass. However, the MIND diet could have improved the anthropometric parameters more in comparison with the control group. The within-group comparison analysis showed that these improvements were found over time in both the MIND and the control group (Table 3), which expressed the effect of hypocaloric dietary pattern on anthropometric indices.

**Table 3.** Changes in anthropometric parameters after three months follow up in the MIND diet group versus the control group.

| *Anthropometric parameters* | *MIND group* |  | *Control group* |  | *F<sup>a</sup>*<br>(1,35) | *Eta<sup>β</sup>* | *P value<sup>ε</sup>* |
| --- | --- | --- | --- | --- | --- | --- | --- |
|  | *Mean changes* | *P value<sup>ε</sup>* | *Mean changes* | *P value* |  |  |  |
|  | (*±SE*) |  | (*±SE*) |  |  |  |  |
| ***TBW (L)*** | 1.71±0.68 | **0.020** | 1.30±0.44 | 0.011 | 0.21 | 0.006 | 0.647 |
| ***PBF (%)*** | -5.16±0.82 | **<0.001** | -2.56±0.72 | 0.003 | 4.96 | 0.124 | **0.032** |
| ***FFM (kg)*** | 2.54±0.52 | **<0.001** | 1.95±0.64 | 0.009 | 0.49 | 0.014 | 0.486 |
| ***BMI (kg/m<sup>2</sup>)*** | -1.55±0.11 | **<0.001** | -0.92±0.07 | <0.001 | 16.86 | 0.325 | **<0.001** |
| ***Weight (kg)*** | -3.98±0.29 | **<0.001** | -2.34±0.17 | <0.001 | 18.21 | 0.342 | **<0.001** |
| ***WC (cm)*** | -3.54±0.56 | **<0.001** | -1.46±0.49 | 0.011 | 6.74 | 0.162 | **0.014** |
| ***HC (cm)*** | -2.27±0.23 | **<0.001** | -0.30±0.12 | 0.009 | 37.66 | 0.518 | **<0.001** |

Results from the repeated measure ANOVA for group × time interaction for anthropometrics data after three months follow up. Abbreviation: MIND = Mediterranean-DASH Intervention for Neurodegenerative Delay, TBW= Total Body Water, PBF= Percent of Body Fat, FFM= Fat-Free Mass, BMI=Body Mass Index, WC= Waist Circumferences, HC= Hip Circumferences. <sup>a</sup>=F-value (df1 for the numerator, df2 for the denominator), <sup>β</sup>=Partial eta squared for the ratio of variance associated with an effect, <sup>ε</sup>= *p*-value for differences between groups, <sup>ε</sup>= *p*-value for differences within groups.

**Figure 2.**
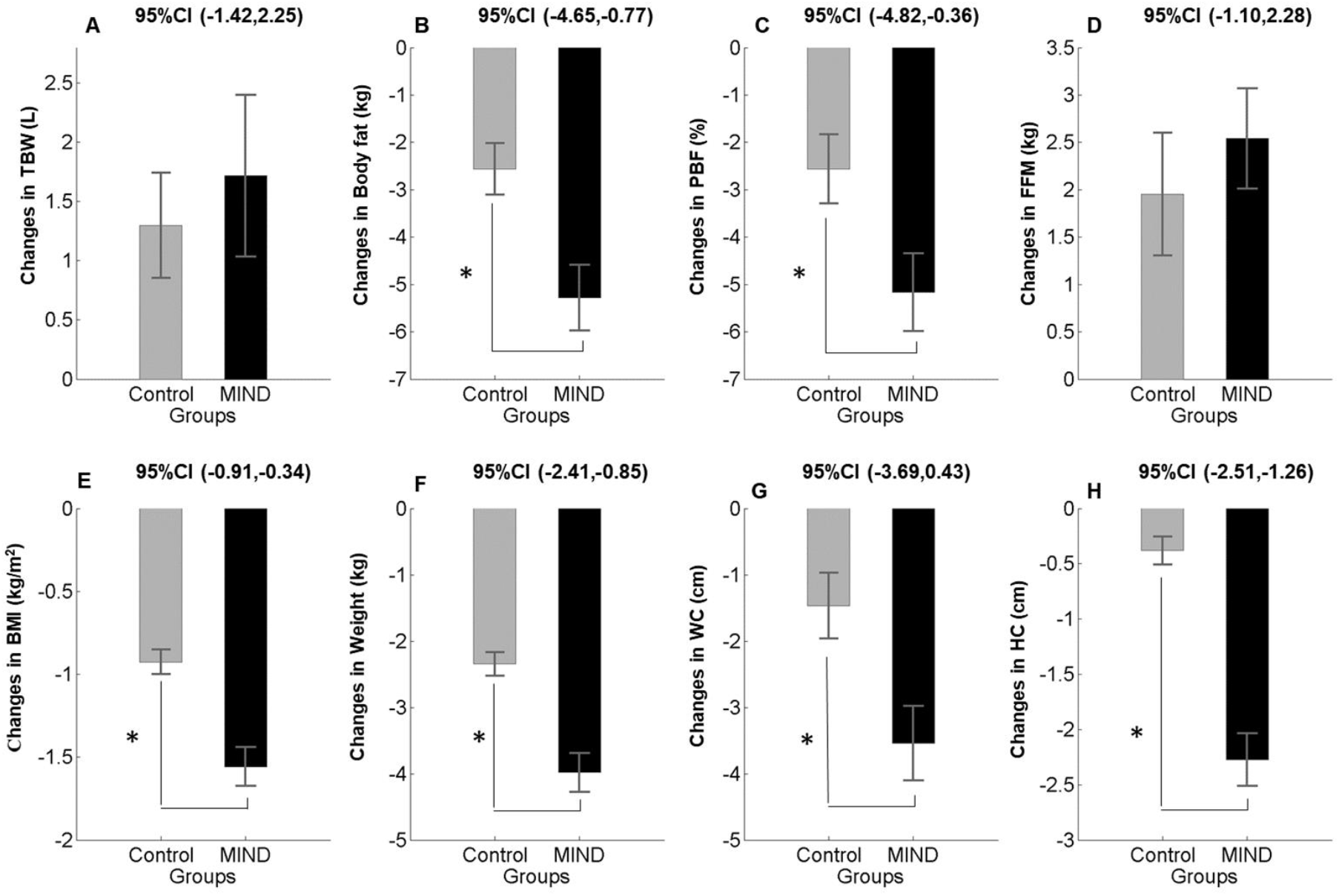
Anthropometric changes (mean and standard error of the mean) in the MIND diet group (black color) and control group (gray color) at baseline and follow up. Note that *P*-value < 0.05 in a repeated measure ANOVA test indicating significant improvement in Body Mass Index, weight, waist circumference (panel E, F, and G) and percent of body fat (panel C) as well as hip circumference (panel H) in MIND diet group in comparison with the control group. Abbreviation: TBW= Total Body Water, PBF= Percent of Body Fat, FFM= Fat-Free Mass, BMI=Body Mass Index, WC= Waist Circumferences, HC= Hip Circumferences.

### 3.4. Changes in Metabolic Profiles

As shown in Table 4, the repeated measure ANOVA for group × time interaction revealed, there was a significant interaction effect for plasma levels of leptin (*p* = 0.035, Figure2, panel A), as well as Ghrelin (*p* =0.02, Figure2, panel B). This effect also found for plasma levels of GLP-1 (*p* = 0.014, Figure2, panel C) too. Indicating that plasma levels of these hunger hormones can be improved better by MIND diet intervention compared to the control group. Except for Leptin, the within-group comparison showed no statistically significant difference in the primary endpoint after three months. Mean differences in each group at each time point presented in Figure 3.

**Table 4.**
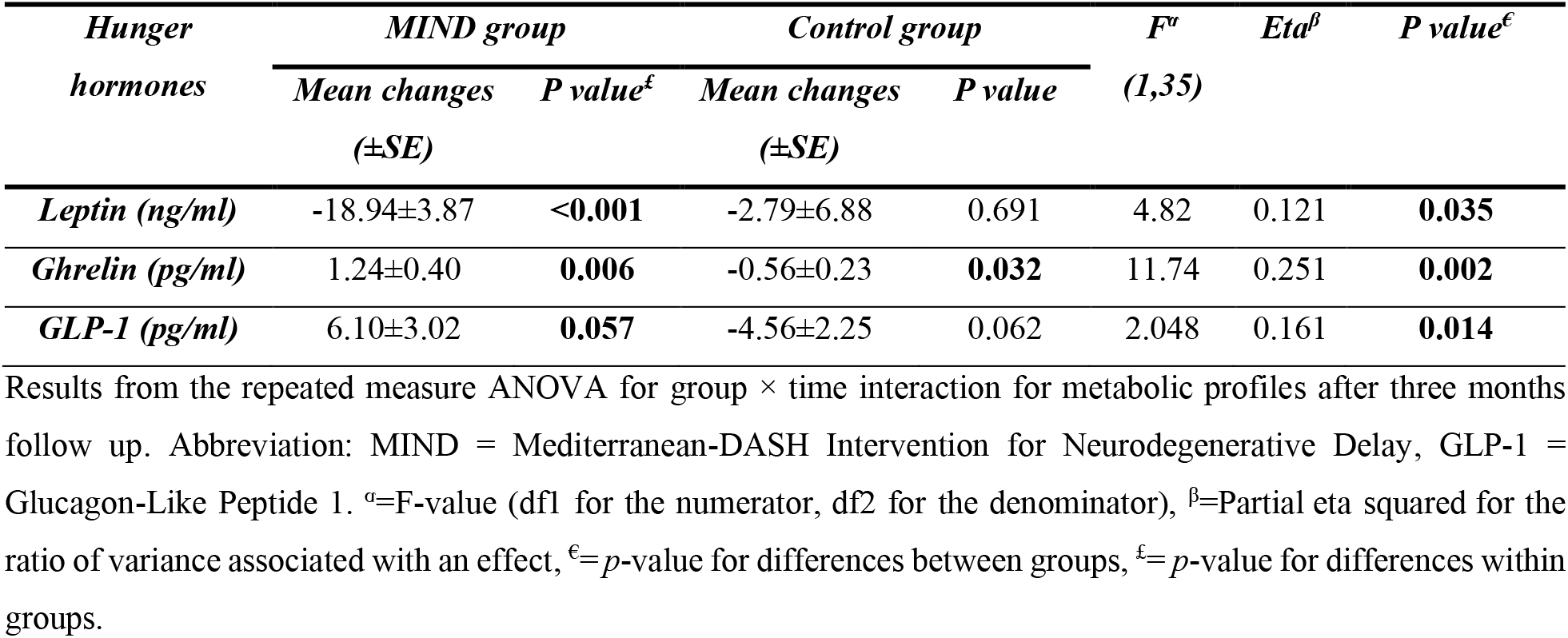
Changes in hunger hormones after three months follow up in the MIND diet group versus the control group.

**Figure 3.**
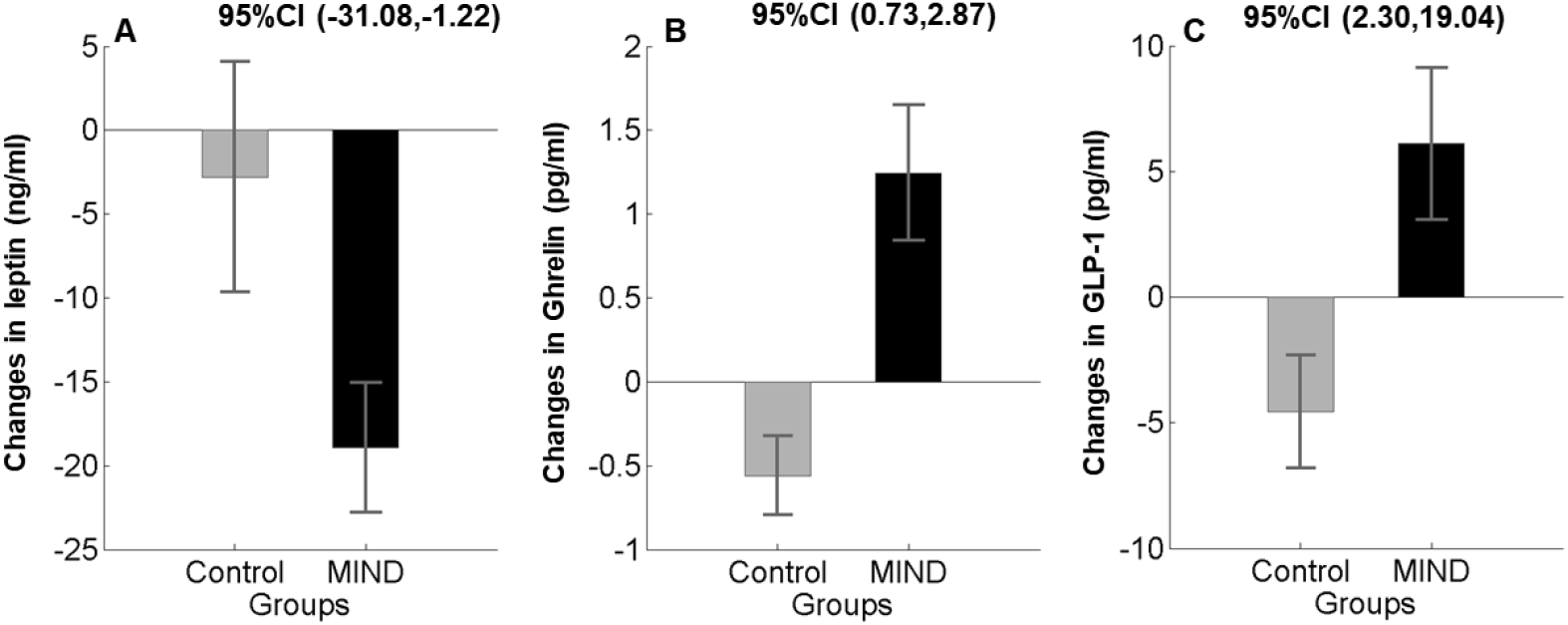
Changes in metabolic profile (mean and standard error of the mean) in the MIND diet group (black color) and control group (gray color) at baseline and follow up. Note that the MIND diet group exhibited improvement in plasma levels of Leptin, ghrelin, and GLP-1 (panel A, B, D, and C) than the control group in a repeated measure group × time interaction ANOVA. Abbreviation: MIND = Mediterranean-DASH Intervention for Neurodegenerative Delay, GLP-1 = Glucagon-Like Peptide 1.

### 3.5. Changes in Brain Structures

In the current study, we first used whole-brain analysis to investigate changes in brain structures, so the results didn’t show significant differences in any of the brain regions between two groups, which might be due to the small sample size for this analysis. To overcome this problem, we anatomically selected areas of the brain that had been responded to the special dietary pattern in previous studies. These regions included fusiform, parahippocampal, medial, superior, and temporal gyrus.

Our results diminished to found any interaction effects for brain structures. Indicated that there were no statistically significant differences between the MIND diet and control group on brain structures after three months. In addition, our result revealed that there were no significant between-group mean differences in brain structure between participants allocated to the MIND diet and the control group.

## 4 Discussion

To our knowledge, this is the first randomized controlled trial to investigate the effect of short-term MIND diet intervention in healthy obese women. For the primary endpoints, we measured anthropometric parameters as well as metabolic profiles. The secondary endpoint was to investigate the effect of the MIND diet on brain structure differences. Our results showed that mean changes in weight, percent of body fat, and waist circumference was more decreased in the MIND diet group. On the other hand, three months of MIND diet intervention improved metabolic profiles in comparison with the control group. We could not find any significant differences in the brain structure of two study groups.

Our analysis indicated a more significant effect of the MIND diet on weight, independent of energy intake, which shows the synergistic effect of the elements that make up the MIND diet along with the calorie restriction. In the present study, BMI in line with the percent of body fat decreased significantly more in the MIND diet group compared with the control group. Additionally, our result showed that adherence to the MIND diet guidelines could maintain total body water and fat-free mass. In this regard, Vincent et al. revealed that adherence to the Mediterranean diet could more decrease the mean difference of BMI −0.3kg/m (*p* < 0.05) in comparison with the low-fat diet [13]. However, the three-month parallel study on high-risk cardiovascular participants couldn’t detect the significant relationship between the Mediterranean diet and changes in anthropometric parameters [14]. In addition, in line with our results, a random-effects meta-analysis of 19 arms stated that a greater effect of the Mediterranean diet on weight independent of energy intake [15]. In studies conducted on the DASH diet, it was shown that the impact of the DASH diet on body weight was greater (−2.27kg) when accompanied with energy-restricted diets for short term periods [9]. A possible correlation has been suggested, which is, the MIND diet is based on the Mediterranean and DASH dietary patterns. Therefore, part of its positive effects on anthropometric parameters is due to the effects of mentioned patterns on weight loss.

By Looking at the functional mechanism of the MIND dietary pattern, it could be concluded that this new pattern has taken steps to enrich two old dietary patterns (the Mediterranean and DASH diet) in promoting general and especially brain health. In this line, observational studies revealed that high MUFA olive oil content of the diet was associated with a decreasing obesity rate [16]. Also, prospective studies declare that adherence to the olive oil reach diet is inversely related to abdominal obesity but not BMI [17]. One of the possible mechanisms in this regard is that the oleic acid in olive oil activates oleoylethanolamide, which is one of the intestinal satiety messengers in the brain [18]. On the other hand, the preventive properties of berries and anthocyanins against body fat accumulation was first reported in 2003 [19]. The authors demonstrated that obesity induced by a high-fat diet could be significantly suppressed by berries extract in C57BL/6J mice.

Regarding markers of inflammation, our findings showed that adherence to the MIND diet was associated with lower levels of Leptin. Previous studies demonstrated that these changes in plasma levels of Leptin are only noticeable when it was associated with weight loss [20]. This result is consistent with a number of studies revealed that higher adherence to the Mediterranean dietary pattern is directly associated with a significant reduction in plasma levels of Leptin [21]. MIND diet with weight loss significantly reduced plasma leptin levels. It is well known that Leptin is one of the adipokines which are synthesized by adipocytes in proportion to the volume of fat and also play an essential role in the regulation of appetite. In addition to the position of Leptin as a satiety factor, it is also called a considerable molecule at the cornerstone of metabolism, inflammation, and neurodegeneration [22]. Our results also point to an increase in plasma levels of ghrelin. This result is in line with clinical trial studies that showed dietary-induced weight loss triggers a significant reduction in ghrelin level, directly proportional to the BMI reduction [23]. Ghrelin is a peptide hormone, primarily produced by specialized cells of the stomach, and similarly to Leptin, it also has a vital role in the regulation of metabolism [24].

Our result demonstrated that MIND diet intervention could increase the mean change of GLP-1 secretion in three months. However, we found a decrease in the secretion in the control group. The role of GLP-1 in the progress of obesity has been suggested in part because of the physiological effects of the hormone on appetite and food intake, and studies have declared that GLP-1 secretion may be decreased in obesity [25]. In extensive research on diabetes and non-diabetics participants, it was shown that body mass index (BMI) as an important factor has a significant negative relationship with GLP-1 secretion [26]. Besides, another study found that the secretion of incretin, which stimulates insulin secretion, is inversely related to glucose tolerance and BMI [27]. This association is independent and growing. According to the findings of our study, there might be other reasons, including food pattern components, other than calorie restriction alone, which could affect plasma levels of GLP-1.

Additionally, consumption of green leafy vegetables, as well as whole grains, offer anti-inflammatory and anti-oxidative effects of the MIND diet, which can have a detrimental impact on serum levels of pro-inflammatory factors such as Leptin and ghrelin [28]. Finally, low amounts of red meat and saturated fatty acid content of the MIND diet lead to an improvement in body weight, glucose metabolism, and cardiovascular risk factors [29]. In this regard, the MIND diet favors weight loss for reasons such as plenty of dietary fibers, low energy density, low glycemic load, and high-water content. Therefore, the synergistic effect of MIND diet components might have a pivotal effect on anthropometric parameters and metabolic profile of obese participants.

Results of our trial demonstrated that participants in the MIND diet group din not differed in some brain regions than participants in the control group after three months. Our findings inconsistent with Lauri et al. This study showed that calorie restriction could partially reverse white matter expansion in obese participants. However, their results could not find any significant differences in regional gray matter volume after dieting [30]. This RCT was performed on 16 obese subjects continued a very low-calorie diet for 6week. One of the possible mechanisms is the terms of calorie restriction. In our trial, we used at least 1500 kcal/d against a very low-calorie diet in Lauri’s study. The majority of the studies with significant effects conducted in a longer duration, which is one of the limitations of our research.

On the other hand, the results of animal models’ studies show that about 5% of unesterified plasma fatty acids enter the animal’s brain through cerebral blood flow. Obesity is associated with the elevation of free fatty acids in the plasma, along with the accumulation of fatty acids in fat cells as well as in the brain [31]. Therefore, dietary patterns and calorie restriction could reverse, thus buildup. It was beyond the scope of our study to investigate this relationship in obese and lean subjects.

### 4.1. Strengths and limitations

Our trial has several strengths. First, to our knowledge, it was the first randomized trial to investigate the effect of MIND intervention in healthy obese women. Second, the present study was a well-controlled dietary intervention trial in which very high compliance was obtained. Third, both study groups guided to the calorie-restricted diet to compare between the MIND diet and calorie restriction separate from each other.

However, our study also has limitations. The current trial was a single-center, controlled recruitment study, which examined a sample of well-educated healthy volunteers. So, we cannot generalize our data to a big part of society. Besides, the short length of our study, along with the small sample size, can be the other limitation.

### 4.2. Conclusion

In summary, the result of our trial for the first time showed that using the MIND diet intervention in healthy obese women can improve anthropometric parameters as well as metabolic profiles. This effect could be strengthened by the calorie restriction too. Although the results of our study have failed to find a link between brain structure and the MIND dietary pattern. Exploring the validity of our findings in future multicenter studies with larger sample size and longer duration will be necessary.

## Data Availability

All data referred to in the manuscript.

## Conflict of interest statement

The authors declare no conflict of interest.

### Acknowledgments

The authors would like to thank the participants for their kind and enthusiastic corporation. This trial was financially supported by Shiraz University of Medical Science, Shiraz, Iran (Grant numbers: 97-01-84-17299).

## Statement of Authorship

Golnaz Arjmand: Conceptualization, Methodology, Data curation, Writing, Formal analysis. Mohammad Hassan Eftekhari: Conceptualization, Methodology, Review, and editing Supervising. Mojtaba Abbas-Zadeh: Methodology, Data curation, Review and editing, Formal analysis. Majid fardaei: Methodology.

## References

1. Heymsfield, S.B. and T.A. Wadden, Mechanisms, pathophysiology, and management of obesity. New England Journal of Medicine, 2017. 376(3): p. 254–266.

2. Cui, H., M. López, and K. Rahmouni, The cellular and molecular bases of leptin and ghrelin resistance in obesity. Nature Reviews Endocrinology, 2017. 13(6): p. 338.

3. Sánchez-Garrido, M.A., et al., GLP-1/glucagon receptor co-agonism for treatment of obesity. Diabetologia, 2017. 60(10): p. 1851–1861.

4. Verstynen, T.D., et al., Increased body mass index is associated with a global and distributed decrease in white matter microstructural integrity. Psychosomatic medicine, 2012. 74(7): p. 682.

5. Pannacciulli, N., et al., Brain abnormalities in human obesity: a voxel-based morphometric study. Neuroimage, 2006. 31(4): p. 1419–1425.

6. Taki, Y., et al., Relationship between body mass index and gray matter volume in 1,428 healthy individuals. Obesity, 2008. 16(1): p. 119–124.

7. Rullmann, M., et al., Gastric-bypass surgery induced widespread neural plasticity of the obese human brain. Neuroimage, 2018. 172: p. 853–863.

8. Varady, K., S. Bhutani, and C. Braunschweig, Degree of weight loss required to improve adipokine concentrations and decrease fat cell size in severely obese women. Metabolism: clinical and experimental, 2009. 58: p. 1096–101.

9. Soltani, S., et al., The effect of dietary approaches to stop hypertension (DASH) diet on weight and body composition in adults: a systematic review and meta-analysis of randomized controlled clinical trials. Obesity reviews, 2016. 17(5): p. 442–454.

10. Morris, M.C., et al., MIND diet slows cognitive decline with aging. Alzheimer’s & dementia, 2015. 11(9): p. 1015–1022.

11. Mahan, L.K. and J.L. Raymond, Krause’s Food & the Nutrition Care Process, Mea Edition E-Book. 2016: Elsevier.

12. Ségonne, F., et al., A hybrid approach to the skull stripping problem in MRI. Neuroimage, 2004. 22(3): p. 1060–1075.

13. Vincent-Baudry, S., et al., The Medi-RIVAGE study: reduction of cardiovascular disease risk factors after a 3-mo intervention with a Mediterranean-type diet or a low-fat diet. The American journal of clinical nutrition, 2005. 82(5): p. 964–971.

14. Estruch, R., et al., Effects of a Mediterranean-style diet on cardiovascular risk factors: a randomized trial. Annals of internal medicine, 2006. 145(1): p. 1–11.

15. Esposito, K., et al., Mediterranean diet and weight loss: meta-analysis of randomized controlled trials. Metabolic syndrome and related disorders, 2011. 9(1): p. 1–12.

16. Pérez-Martínez, P., et al., Mediterranean diet rich in olive oil and obesity, metabolic syndrome and diabetes mellitus. Current pharmaceutical design, 2011. 17(8): p. 769–777.

17. Romaguera, D., et al., Adherence to the Mediterranean diet is associated with lower abdominal adiposity in European men and women. The Journal of nutrition, 2009. 139(9): p. 1728–1737.

18. López-Miranda, J., et al., Olive oil and health: summary of the II international conference on olive oil and health consensus report, Jaén and Córdoba (Spain) 2008. Nutrition, metabolism and cardiovascular diseases, 2010. 20(4): p. 284–294.

19. Tsuda, T., et al., Dietary cyanidin 3-O-β-D-glucoside-rich purple corn color prevents obesity and ameliorates hyperglycemia in mice. The Journal of nutrition, 2003. 133(7): p. 2125–2130.

20. Rashad, N.M., et al., Effect of a 24-week weight management program on serum leptin level in correlation to anthropometric measures in obese female: A randomized controlled clinical trial. Diabetes & Metabolic Syndrome: Clinical Research & Reviews, 2019. 13(3): p. 2230–2235.

21. Sureda, A., et al., Adherence to the mediterranean diet and inflammatory markers. Nutrients, 2018. 10(1): p. 62.

22. Park, H.-K. and R.S. Ahima, Physiology of leptin: energy homeostasis, neuroendocrine function and metabolism. Metabolism, 2015. 64(1): p. 24–34.

23. Kotidis, E.V., et al., Serum ghrelin, leptin and adiponectin levels before and after weight loss: comparison of three methods of treatment–a prospective study. Obesity surgery, 2006. 16(11): p. 1425–1432.

24. Mani, B.K. and J.M. Zigman, Ghrelin as a survival hormone. Trends in Endocrinology & Metabolism, 2017. 28(12): p. 843–854.

25. Heppner, K.M. and D. Perez-Tilve, GLP-1 based therapeutics: simultaneously combating T2DM and obesity. Frontiers in neuroscience, 2015. 9: p. 92.

26. Vigneshwaran, B., et al., Impact of sleeve gastrectomy on type 2 diabetes mellitus, gastric emptying time, glucagon-like peptide 1 (GLP-1), ghrelin and leptin in non-morbidly obese subjects with BMI 30–35.0 kg/m 2: a prospective study. Obesity surgery, 2016. 26(12): p. 2817–2823.

27. Muscelli, E., et al., Separate impact of obesity and glucose tolerance on the incretin effect in normal subjects and type 2 diabetic patients. Diabetes, 2008. 57(5): p. 1340–1348.

28. Maruyama, C., et al., Effects of green-leafy vegetable intake on postprandial glycemic and lipidemic responses and α-tocopherol concentration in normal weight and obese men. Journal of nutritional science and vitaminology, 2013. 59(4): p. 264–271.

29. Montonen, J., et al., Consumption of red meat and whole-grain bread in relation to biomarkers of obesity, inflammation, glucose metabolism and oxidative stress. European journal of nutrition, 2013. 52(1): p. 337–345.

30. Haltia, L.T., et al., Brain white matter expansion in human obesity and the recovering effect of dieting. The Journal of Clinical Endocrinology & Metabolism, 2007. 92(8): p. 3278–3284.

31. Rapoport, S.I., In vivo fatty acid incorporation into brain phosholipids in relation to plasma availability, signal transduction and membrane remodeling. Journal of Molecular Neuroscience, 2001. 16(2-3): p. 243–261.

